# Oxylipins and the Surgical Classification of Chronic Thromboembolic Pulmonary Hypertension

**DOI:** 10.1101/2022.05.20.22275399

**Authors:** Mona Alotaibi, Timothy Fernandes, Amber Tang, Kim Kerr, Timothy Morris, Atul Malhotra, Jason X-Y. Yuan, Victor Pretorius, Michael Madani, Jeramie D. Watrous, Tao Long, Michael W. Pauciulo, William C. Nicholls, Mohit Jain, Susan Cheng, Nick Kim

## Abstract

Surgically accessible lesions of chronic thromboembolic pulmonary hypertension (CTEPH) are classified as proximal or distal based on the distribution of thrombus burden in the pulmonary vasculature post operatively. Surgically accessible distal CTEPH lesions typically has a higher risk profile and worse clinical outcomes in less experienced centers, but the underlying molecular differences between proximal and distal CTEPH lesions remain unknown. Oxylipins, a diverse group of bioactive lipid mediators, have previously been implicated in a range of disorders including pulmonary hypertension. Therefore, we sought to characterize oxylipin profiles among patients with proximal and distal operable CTEPH lesions, as well as those with idiopathic pulmonary arterial hypertension (IPAH). We studied 271 patients with proximal operable CTEPH (n=123), distal operable CTEPH (n=74), or IPAH (n=74). Liquid chromatography-mass spectrometry was used to analyze oxylipin profiles in each patient. We found that patients with distal operable CTEPH had elevated levels of proinflammatory oxylipins while those with proximal CTEPH had an increase in procoagulant oxylipins. Notably, the proinflammatory oxylipins elevated in distal operable CTEPH were similarly elevated in IPAH. These findings suggest that distal operable and proximal CTEPH represent heterogenous disease processes. Furthermore, oxylipin profiles may be useful for potential risk stratification and therapeutic targeting in CTEPH.

## BACKGROUND

Chronic thromboembolic pulmonary hypertension (CTEPH) is a life threatening but treatable cause of pulmonary hypertension associated with venothromboembolism.^1^ Pulmonary hypertension in CTEPH is caused by mechanical obstruction from chronic unresolved thrombi as well as varying degrees of small vessel arteriopathy similar to lesions found in pulmonary arterial hypertension (PAH). Pulmonary endarterectomy (PEA) is the treatment of choice for CTEPH and aims to remove the mechanical obstructive component responsible for PH.^2^ Long-term outcomes in CTEPH are best with PEA. ^3^ Surgical classification of CTEPH can only be determined after PEA and is based on location of the thromboembolic disease found at the time of endarterectomy.^2^ In expert centers, levels III and IV are still deemed operable but pose technical challenges requiring multiple planes of endarterectomy to achieve adequate clearance. Patients predicted to have such distal operable lesions may be turned down for surgery at less experienced surgical centers in lieu of PH targeted medical therapy and/or balloon pulmonary angioplasty (BPA).

Relatively little is known about distinguishing molecular features associated with the level of surgical disease found and removed at the time of PEA. Some studies have suggested that oxylipins, bioactive lipid mediators that are products of polyunsaturated fatty acids, may play a role in the pathogenesis of a multitude of inflammatory conditions with potential risk stratification or therapeutic applications.^4^ Oxylipins have been previously implicated in disease processes, where they can have inflammatory, vasoconstrictive, and proliferative effects.^5^ These compounds have been associated with vascular remodeling in both PAH^6,7^ and CTEPH.^8^ However, to our knowledge, no studies to date have explored the potential role of oxylipin mediators underlying different surgical lesions in CTEPH. As a result, we examined differences in oxylipin profiles among patients with operable proximal and distal CTEPH lesions as well as idiopathic PAH (IPAH).

## METHODS

### Study Cohorts

Patients undergoing bilateral PEA at UCSD from January 2015 to December 2018 were included in the study. Sex and age matched IPAH patients (within 5 years), were included from the PAH biobank. All patients had plasma samples stored in the University of California San Diego biobank (CTEPH) or National Biological Sample and Data Repository for Pulmonary Arterial Hypertension biobank (PAHB). For all patients, pulmonary hypertension diagnosis was confirmed by standard criteria on right heart catheterization.^9^ CTEPH was confirmed based on results of ventilation/perfusion scan and computed tomography scan or pulmonary angiography performed ≥3 months after anticoagulation therapy.^1^ Criterial for PEA and surgical procedures were based on previously published guidelines.^2,10^ Diagnosis of IPAH was established on the prevailing diagnostic criteria for PH at the time of cohort recruitment.^9^ The UCSD IRB approved the study protocols and all patients signed an informed consent at their respective institutions.

### PEA Surgical Specimens

PEA surgery was carried out as described previously.^10,11^ The UCSD surgical classification system^10^ was used to describe the four major levels of CTEPH specimens based on the location of the disease. UCSD level I was defined as lesions starting in the main pulmonary arteries; level II was defined as lesions starting at the level of the lobar or intermediate pulmonary arteries; level III was defined as lesions starting at the level of segmental arteries only; and level IV was defined as lesions starting at the subsegmental branches only. We defined proximal lesions as bilateral UCSD level I/II and distal lesions as bilateral UCSD levels III/IV. If there was a discrepancy in CTEPH level between left and right specimen, the sample was excluded from the analysis. (**Figure 1**).

**Figure.**
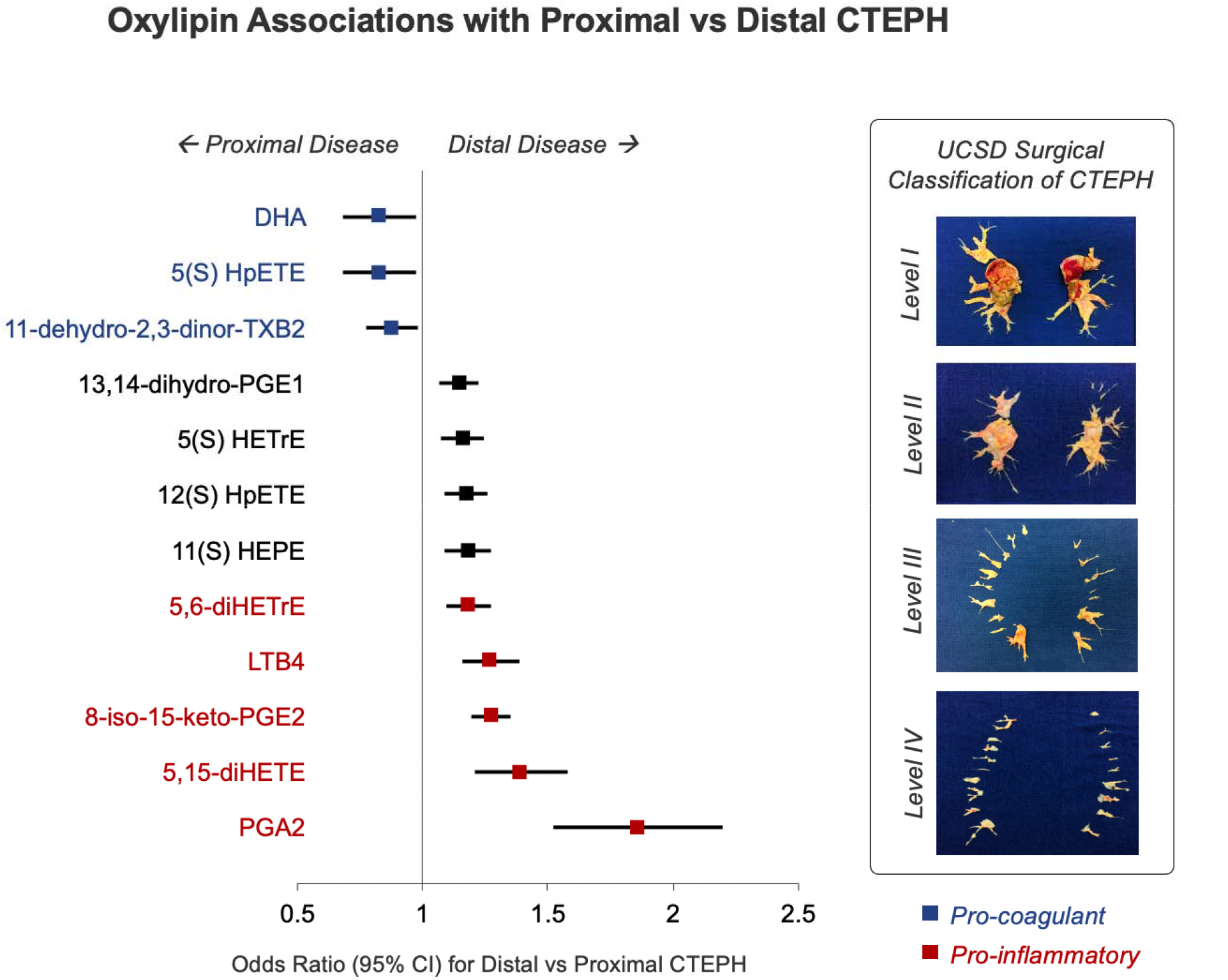
Oxylipin Associations with CTEPH Surgical Lesions. Surgical classification of distal vs proximal CTEPH is based on direct surgical phenotypic assessment of thrombus burden distribution (*inset*). In multivariable analyses adjusting for age, sex, and body mass index, we observed a predominance of inflammatory oxylipins significantly associated with distal versus proximal CTEPH phenotypes. (*Blue markers denote oxylipins implicated in pro-coagulant pathways. Red markers denote oxylipins implicated in inflammatory pathways*.)

### Oxylipins Analysis

For all patients, plasma samples underwent LC-MS based oxylipin profiling using methods we have described previously.^12^ Following Qc/Qa analysis, data were extracted using image processing and machine learning based spectral optimization,^12^ then normalized, aligned, and filtered for statistical analyses.

### Statistical Analysis

Prior to all statistical analyses, oxylipin analyte values were natural logarithmically transformed, then standardized (mean=0, SD=1) to facilitate inter-analyte comparisons. Between-group comparisons were performed using t-test or the Mann-Whitney test for continuous variables and the χ^2^ test for categorical variables. We used logistic regression analysis to identify oxylipins associated with subsegmental versus segmental CTEPH in models adjusting for age, sex, and body mass index (BMI). We then repeated analyses to compare oxylipins levels in subsegmental CTEPH versus IPAH. We considered a Bonferroni corrected *P* value significance threshold of 0.05 divided by a conservative estimate of the total number of unique small molecules (i.e., *p*<10^−3^). All statistical analyses were performed using R v3.5.1.

## RESULTS

Out of 288 CTEPH patients who underwent PEA and had plasma samples stored, 91 were excluded for unmatched surgical levels. A total of n=271 patients with confirmed post-operative distal CTEPH lesions (bilateral levels III/IV, n=74), proximal CTEPH lesions (bilateral level I/II, n=123), or IPAH (n=74) were included in the analysis. Of the 197 patients in our cohort with confirmed CTEPH, 123 had proximal CTEPH and were 57.9% female with an average age of 55 and predominately functional class 3 (**Table 1**). Similarly, those with distal CTEPH (n=74) had a mean age of 52 years and were also predominately female (66.7%) and functional class 3. We observed similar between-group characteristics for patients with proximal versus distal CTEPH patients, including pre-operative hemodynamic profile and prevalence of comorbidities. Furthermore, there were no significant differences in hospital outcomes, including number of days on a ventilator, intensive care unit length of stay, and total length of stay. Both the proximal and distal CTEPH patients experienced significant hemodynamic improvement, although a higher post-operative PVR (3.02 vs 2.41, p-value <0.001) was note din the distal group. Patients with distal CTEPH had longer circulatory arrest time and higher incidence of post-operative reperfusion injury compared to proximal CTEPH (24.3% vs 11.5%, p-value 0.03). (**Tables 1 & 2**).

**Table 1.** Patients’ characteristics.

|  | **Distal** | **Proximal** | ***p*** |
| --- | --- | --- | --- |
| **n** | **74** | **123** |  |
| **Age, years** | 52.30 (16.18) | 55.08 (13.31) | 0.19 |
| **Female** | 38 (66.7) | 55 (57.9) | 0.37 |
| **Body mass index, kg/m2** | 29.90 (8.01) | 30.47 (6.69) | 0.59 |
| **Pulmonary hypertension therapy, (%)** | 44 (59.5) | 64 (52.0) | 0.39 |
| **Endothelin receptor antagonists** | 11 (14.9) | 9 (7.3) | 0.15 |
| **Phosphodiesterase 5 inhibitors** | 10 (13.5) | 17 (13.8) | 1.00 |
| **Prostaglandins** | 5 (6.8) | 6 (4.9) | 0.81 |
| **Riociguat** | 30 (40.5) | 43 (35.0) | 0.53 |
| **Inhaled prostaglandin therapies** | 2 (2.7) | 3 (2.4) | 1.00 |
| **Functional Class** |  |  | 0.76 |
| **1** | 0 (0.0) | 1 (0.8) |  |
| **2** | 20 (27.0) | 32 (26.0) |  |
| **3** | 50 (67.6) | 86 (69.9) |  |
| **4** | 4 (5.4) | 4 (3.3) |  |
| **Coronary artery bypass graft (%)** | 5 (6.8) | 14 (11.4) | 0.42 |
| **Valve repair (%)** | 0 (0.0) | 5 (4.1) | 0.20 |
| **Patent foramen ovale closure (%)** | 17 (23.0) | 28 (22.8) | 1.00 |
| **Circulatory arrest time: Right, minutes** | 32.19 (12.17) | 21.60 (7.71) | <0.001 |
| **Circulatory arrest time: Left, minutes** | 25.42 (10.12) | 19.20 (7.92) | <0.001 |
| **Total circulatory arrest time, minutes** | 57.73 (20.62) | 40.83 (13.99) | <0.001 |
| **Total cardiopulmonary bypass, minutes** | 249.05<br>(38.16) | 252.00<br>(37.84) | 0.60 |
| **Pulmonary thromboendarterectomy: Level Right, (%)** |  |  | <0.001 |
| **0** | 0 (0.0) | 1 (0.8) |  |
| **1** | 0 (0.0) | 36 (29.3) |  |
| **2** | 0 (0.0) | 86 (69.9) |  |
| **3** | 59 (79.8) | 0 (0.0) |  |
| **4** | 15 (20.3) | 0 (0.0) |  |
| **Pulmonary thromboendarterectomy: Level Left, (%)** |  |  | <0.001 |
| **0** | 0 (0.0) | 3 (2.4) |  |
| **1** | 0 (0.0) | 21 (17.1) |  |
| **2** | 3 (4.1) | 99 (80.5) |  |
| **3** | 45 (60.9) | 0 (0.0) |  |
| **4** | 26 (35.1) | 0 (0.0) |  |
| **Mechanical ventilation, days** | 2.33 (2.89) | 3.34 (5.61) | 0.20 |
| **Intensive care unit length-of-stay, days** | 5.08 (4.11) | 5.43 (4.90) | 0.65 |
| **Postoperative length-of-stay, days** | 11.28 (5.72) | 10.69 (5.01) | 0.51 |
| **Reperfusion injury, (%)** | 18 (24.3) | 14 (11.5) | 0.03 |

*Values are reported as mean and standard deviation unless indicated otherwise.*
*Abbreviations:* PH: pulmonary hypertension.

**Table 2.**
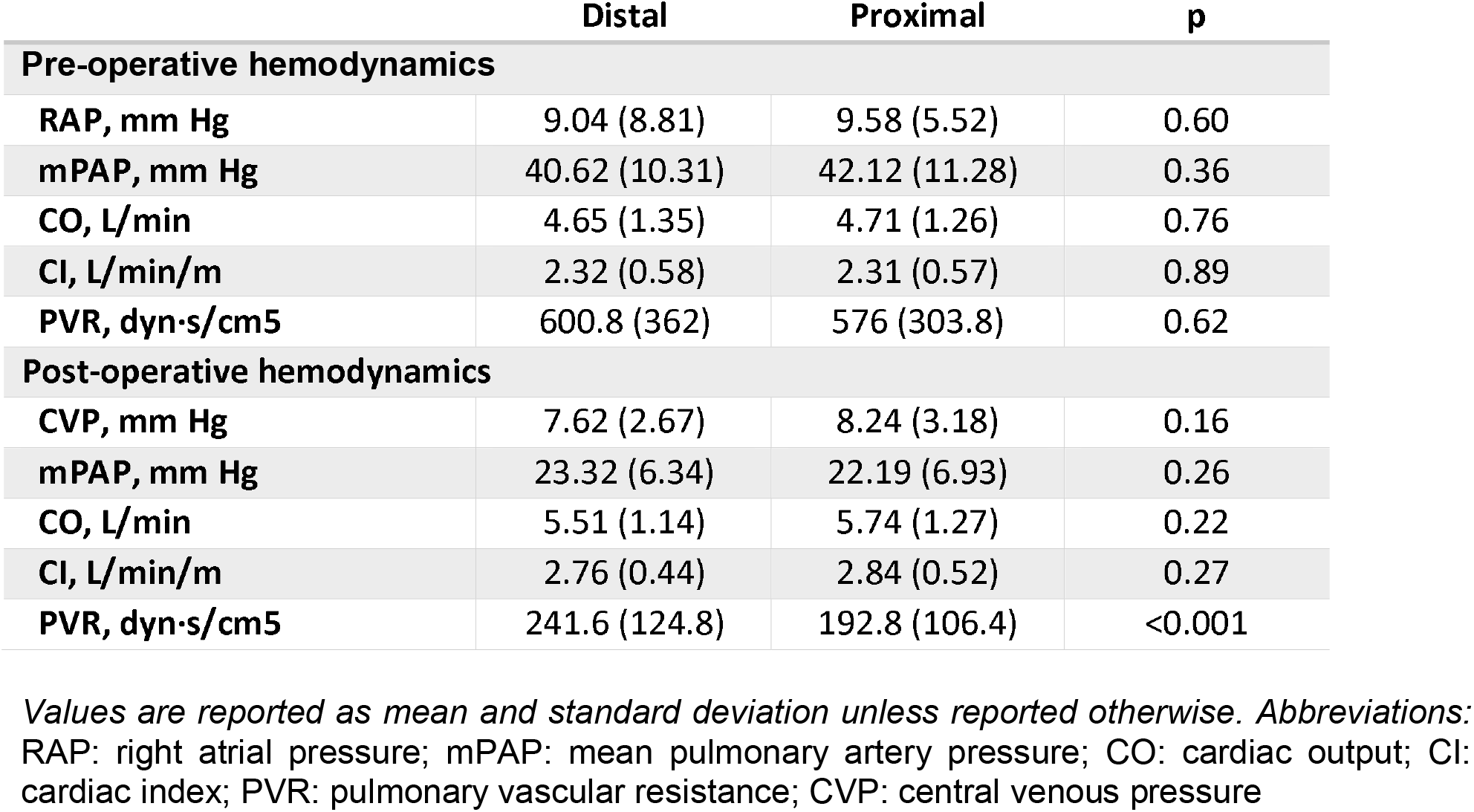
Hemodynamic parameters assessed pre- and post-pulmonary endarterectomy.

Of the 90 oxylipin analytes assayed, we found that 9 oxylipins were increased and 3 oxylipins were decreased in distal compared to proximal CTEPH after adjusting for age, sex, and BMI (**Figure**). Notably, 5 of the 9 oxylipins increased in distal CTEPH have been predominantly implicated in pro-inflammatory pathways (**Figure:** odds ratios ranged from 1.29 to 1.86 per 1-SD log-analyte, P values ranged from 2.6E-09 to 4.4E-04), and all 3 of the oxylipins decreased in distal CTEPH (i.e. increased in proximal CTEPH) have been predominantly implicated in pro-coagulant pathways. Intriguingly, levels of the five inflammatory oxylipins were not significantly different in distal CTEPH compared to IPAH.

## DISCUSSION

Our molecular analyses of bioactive lipids indicate that operable distal CTEPH disease is characterized by an increase in proinflammatory oxylipins, whereas proximal CTEPH is characterized by an increase in procoagulant oxylipins. Despite similar pre-operative hemodynamics, we also found that the oxylipin profile of operable distal CTEPH was more similar to that of IPAH than that of proximal CTEPH. These findings may help identify potentially important molecular targets for future risk assessment and therapeutic applications.

These results extend from previous studies that have identified an important role for oxylipins in pulmonary hypertension vascular remodeling.^6^ Upregulation of 15-HETE synthesis was associated with hypoxic pulmonary hypertension and arterial vasoconstriction. Additionally, 15-HETE has been found to result in increased collagen deposition and wall thickening in patients with pulmonary hypertension.^7^ Comparatively little is known about the role of oxylipins in CTEPH specifically. Furthermore, while CTEPH has been recognized as a heterogenous disease with respect to thrombus distribution and surgical pathological lesions, the molecular mechanisms associated with such heterogeneity have remained unclear.

The predominance of proinflammatory oxylipins in our patient cohort with post-operative distal CTEPH is consistent with the higher risk profile and worse outcomes associated with this population compared to proximal CTEPH. Notably, we found similar oxylipin profiles among patients with post-operative distal CTEPH and those with IPAH. Histologically, CTEPH lesions display diverse pathological changes ranging from irregular intimal thickening to more advance vessels remodeling, including plexiform lesions characteristically seen in IPAH.^13^ Given the diversity of CTEPH pathologies, oxylipin profiles among patients with CTEPH may have pertinent implications for risk stratification in the future.

The cross-sectional design of our study limits its ability to detect temporal associations between oxylipins and clinical characteristics in those with post-operative proximal and distal CTEPH. Generalizability may also be limited as CTEPH patients were from a single institution. Additional studies are needed to further characterize how varying oxylipin profiles may impact pathogenesis of CTEPH. The exact molecular pathways in which the oxylipins identified in our study impact proximal and distal CTEPH also require further elucidation. Nevertheless, our study is unique in its identification of a broad array of oxylipins as well as the relatively large number of patients included with confirmed CTEPH.

Taken together, our findings suggest not only that distal and proximal CTEPH lesions identified at the time of surgery may represent different disease entities but that distal CTEPH may have more remodeling explaining the increase in proinflammatory oxylipins seen compared to proximal CTEPH. PEA surgery is still recommended for distal disease in patients deemed operable, but knowledge of oxylipin profiles prior to surgery may be useful for surgical risk assessment and prediction of post-operative hemodynamics. Although our study was derived from among the largest known biorepositories of CTEPH samples, additional investigations are needed to validate our results and prospectively examine implications of our findings for potentially improving prognostication and therapies for CTEPH.

## Data Availability

Requests for de-identified data may be directed to the corresponding authors (NHK, SC) and will be reviewed by the Office of Research Administration at UCSD prior to issuance of data sharing agreements. Data limitations are designed to ensure patient and participant confidentiality.

## Non-standard Abbreviations and Acronyms

BMI: Body mass index
CTEPH: Chronic thromboembolic pulmonary hypertension
IPAH: Idiopathic pulmonary arterial hypertension
LC-MS: Liquid chromatography - mass spectrometry
mRAP: Mean right atrial pressure
PAH: Pulmonary arterial hypertension
PEA: Pulmonary endarterectomy
PVR: Pulmonary vascular resistance
RHC: Right heart catheterization

## ACKNOWLEDGMENTS

We thank contributors who collected samples used in this study, as well as patients and their families, whose help and participation made this work possible.

